# Higher maternal adiposity reduces offspring birth weight if associated with a metabolically favourable profile

**DOI:** 10.1101/2020.05.25.20112441

**Authors:** WD Thompson, RN Beaumont, A Kuang, NM Warrington, Y Ji, J Tyrrell, AR Wood, D Scholtens, BA Knight, DM Evans, WL Lowe, G Santorelli, R Azad, D Mason, AT Hattersley, TM Frayling, H Yaghootkar, MC Borges, DA Lawlor, RM Freathy

**Affiliations:** Institute of Biomedical and Clinical Science, College of Medicine and Health, University of Exeter, Exeter, UK; MRC Integrative Epidemiology Unit, University of Bristol, Bristol, UK; Department of Preventative Medicine, Northwestern University Feinberg School of Medicine, Chicago, IL, USA; University of Queensland Diamantina Institute, University of Queensland, Brisbane, Australia; K.G. Jebsen Center for Genetic Epidemiology, Department of Public Health and Nursing, NTNU, Norwegian University of Science and Technology, Norway; Department of Medicine, Northwestern University Feinberg School of Medicine, Chicago, IL 60611, USA; Bradford Institute for Health Research, Bradford Royal Infirmary, Duckworth Lane, Bradford, UK; Department of Biochemistry, Bradford Royal Infirmary, Bradford, UK; Population Health, Bristol Medical School, University of Bristol, Bristol, UK; Bristol NIHR Biomedical Research Centre, Bristol, UK

**Keywords:** Adiposity, ALSPAC, BiB, BMI, EFSOCH, glucose, HAPO, insulin, Mendelian randomization, UKB

## Abstract

**Aims/Hypothesis:** Higher maternal BMI during pregnancy results in higher offspring birth weight, but it is not known whether this is solely the result of adverse metabolic consequences of higher maternal adiposity, such as maternal insulin resistance and fetal exposure to higher glucose levels, or whether there is any effect of raised adiposity through non-metabolic (e.g. mechanical) factors. We aimed to use genetic variants known to predispose to higher adiposity coupled with a favourable metabolic profile, in a Mendelian Randomisation (MR) study comparing the effect of maternal “metabolically favourable adiposity” on offspring birth weight with the effect of maternal general adiposity (as indexed by BMI).

**Methods:** To test the causal effects of maternal metabolically favourable adiposity or general adiposity on offspring birth weight, we performed two sample MR. We used variants identified in large genetic association studies as associated with either higher adiposity and a favourable metabolic profile, or higher BMI (N = 442,278 and N = 322,154 for metabolically favourable adiposity and BMI, respectively). We then used data from the same variants in a large genetic study of maternal genotype and offspring birth weight independent of fetal genetic effects (N = 406,063 with maternal and/or fetal genotype effect estimates). We used several sensitivity analyses to test the reliability of the results. As secondary analyses, we used data from four cohorts (total N = 9,323 mother-child pairs) to test the effects of maternal metabolically favourable adiposity or BMI on maternal gestational glucose, anthropometric components of birth weight and cord-blood biomarkers.

**Results:** Higher maternal adiposity with a favourable metabolic profile was associated with lower offspring birth weight (−94 (95% CI: −150 to −38) grams per 1 SD (6.5%) higher maternal metabolically favourable adiposity). By contrast, higher maternal BMI was associated with higher offspring birth weight (35 (95% CI: 16 to 53) grams per 1 SD (4 kg/m^2^) higher maternal BMI). Sensitivity analyses were broadly consistent with the main results. There was evidence of outlier SNPs for both exposures and their removal slightly strengthened the metabolically favourable adiposity estimate and made no difference to the BMI estimate. Our secondary analyses found evidence to suggest that maternal metabolically favourable adiposity decreases pregnancy fasting glucose levels whilst maternal BMI increases them.

The effects on neonatal anthropometric traits were consistent with the overall effect on birth weight, but the smaller sample sizes for these analyses meant the effects were imprecisely estimated. We also found evidence to suggest that maternal metabolically favourable adiposity decreases cord-blood leptin whilst maternal BMI increases it.

**Conclusions/Interpretation:** Our results show that higher adiposity in mothers does not necessarily lead to higher offspring birth weight. Higher maternal adiposity can lead to lower offspring birth weight if accompanied by a favourable metabolic profile.

**Research in Context:** *What is already known about this subject?:*

- Studies in non-pregnant people with obesity suggest many people can have a “metabolically healthy” form of obesity, but effects in pregnancy and on offspring are not known.
- Multiple lines of evidence show that higher maternal BMI is causally associated with higher offspring birth weight, and that this may be mediated by the fetal insulin response to higher maternal gestational glucose.
- Recently, genetic variants have been identified, where one allele is associated with higher adiposity but lower risk of type II diabetes and favourable metabolic profile, including lower insulin and glucose levels, so called “metabolically favourable adiposity”; the mechanism is thought to be due to greater subcutaneous adipose storage capacity that leads to higher insulin sensitivity.

*What is the key question?:*

- What is the effect of maternal metabolically favourable adiposity on birth weight, and how does it compare with the effect of maternal BMI on birth weight?

*What are the new findings?:*

- Higher maternal adiposity can lead to lower, not higher birth weight, if it is also associated with a metabolically favourable profile; this contrasts with the effect of higher maternal general adiposity (BMI), on higher birth weight.
- Higher maternal metabolically favourable adiposity causes lower maternal fasting plasma glucose levels, most likely due to higher insulin sensitivity; in contrast higher maternal general adiposity leads to higher maternal fasting plasma glucose levels, most likely due to lower insulin sensitivity.

*How might this impact on clinical practice in the foreseeable future?:*

- Identifying ways of stratifying overweight and obese pregnant women into those with and without metabolically favourable adiposity could allow for targeted management to obtain healthy fetal growth and birth weight.

## Introduction

Maternal body mass index (BMI), an index of general adiposity, is strongly associated with higher offspring birth weight in observational studies[1]. Mendelian Randomisation (MR) studies of BMI in pregnant women support these associations as causal[2, 3]. High birth weight is associated with adverse perinatal and neonatal outcomes[4]. It is also positively correlated with BMI in adulthood[5], which is associated with increased risk of type II diabetes and cardiovascular disease[6].

A likely mechanism for the association of higher maternal BMI with higher offspring birth weight is via its adverse metabolic consequences. For example, increased maternal BMI results in increased maternal insulin resistance and consequently raised maternal circulating glucose levels. As glucose crosses the placenta via facilitated diffusion, this results in increased insulin secretion by the fetus (maternal insulin cannot cross the placenta[7]). Insulin acts as a growth hormone, and higher fetal insulin secretion leads to increased fetal skeletal growth and fat deposition, resulting in higher birth weight due to greater fat and lean mass[8].

Common genetic variants have been identified where one allele is associated with higher adiposity yet a lower risk of type II diabetes and favourable levels of other metabolic traits[9]. These “metabolically favourable adiposity” alleles may have these effects because they are associated with increased adiposity in the more metabolically stable subcutaneous adipose tissue and with decreased adiposity in the liver[9]. These variants strongly overlap with those identified by a different approach, which search for alleles associated with higher fasting insulin and triglycerides levels, and lower HDL-cholesterol levels independent of BMI[10] Such alleles define a phenotype of “limited peripheral adipose storage capacity” i.e. the opposite alleles to those we describe here as metabolically “favourable”. It is unknown whether the metabolically favourable adiposity alleles in pregnant women affect offspring birth weight. Lack of a positive association of maternal metabolically favourable adiposity alleles with offspring birth weight would be compatible with the hypothesis that the effect of maternal BMI on birth weight is driven by the metabolic consequences of general adiposity and not by adiposity per se.

Our aim was to determine the effect of metabolically favourable adiposity on birth weight and to compare this with the effect of maternal general adiposity on birth weight. In our primary analysis, we used alleles associated with higher maternal metabolically favourable adiposity as genetic instruments to measure the effect of maternal adiposity on birth weight when coupled with “favourable” metabolic effects, using data from large genome-wide association studies (GWAS)[2, 9]. We hypothesised that higher maternal metabolically favourable adiposity would either not associate with birth weight or would associate with lower birth weight if it resulted in lower maternal circulating glucose levels. We compared any effects of maternal favourable adiposity to those of general adiposity (assessed by BMI). In a secondary (exploratory) study, we used available individual-level data on mothers and babies to explore potential effects of maternal favourable vs general adiposity on birth weight related metabolic (e.g. maternal glucose, cord insulin) and anthropometric (e.g. head circumference, skinfold thickness) traits.

## Methods

The study design and different data sources are summarized in **<u>Figure 1</u>** and **<u>sFigure1</u>**, with **<u>Table 1</u>** and **<u>Table 2</u>**, providing more data on each contributing study. Further details, including details of participant consent and ethics approvals, are described in the **<u>supplementary methods</u>**.

**Figure 1:**
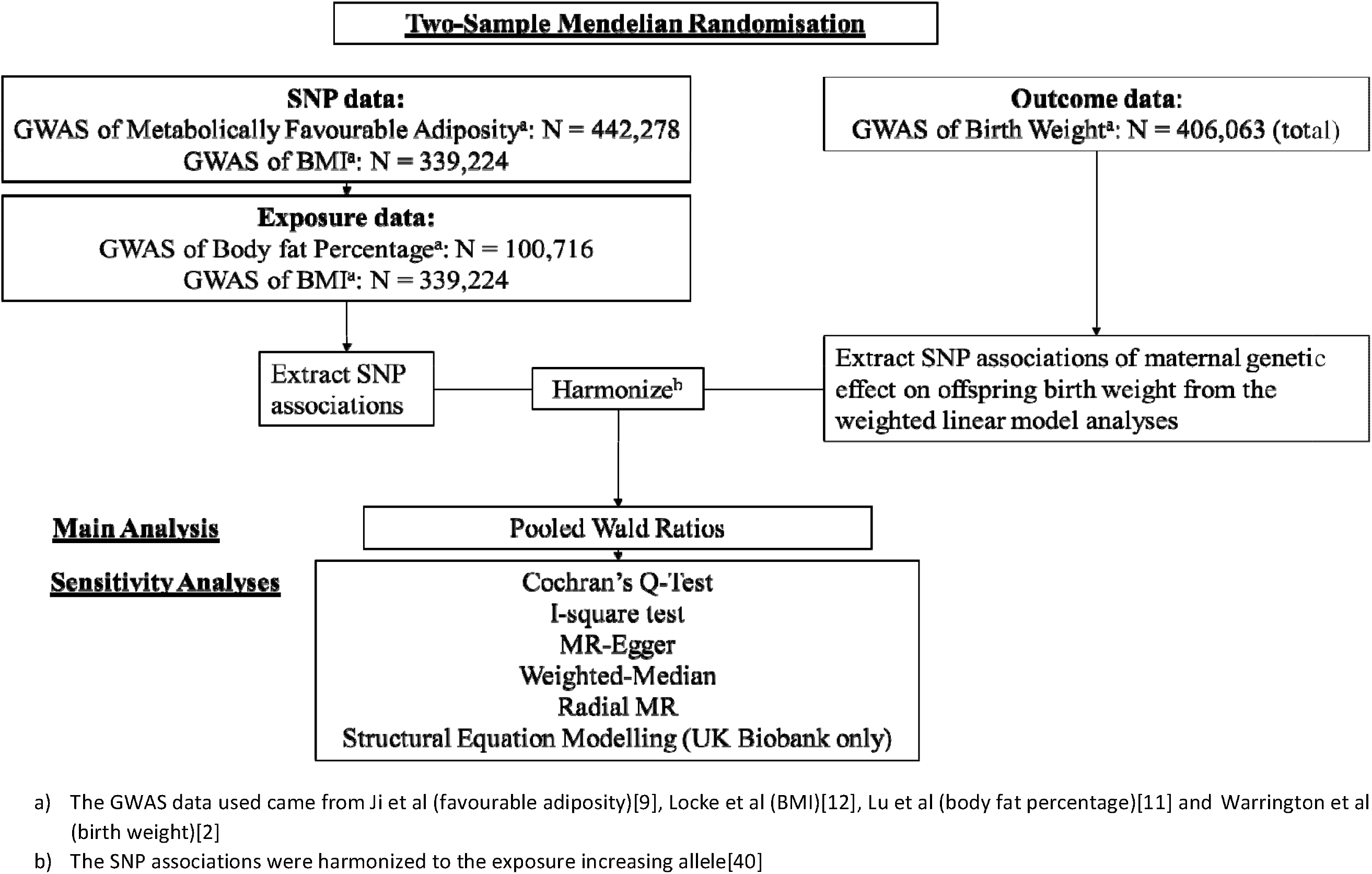
Diagram summarising the key data sources and analysis steps for the primary analyses.

**Table 1:**
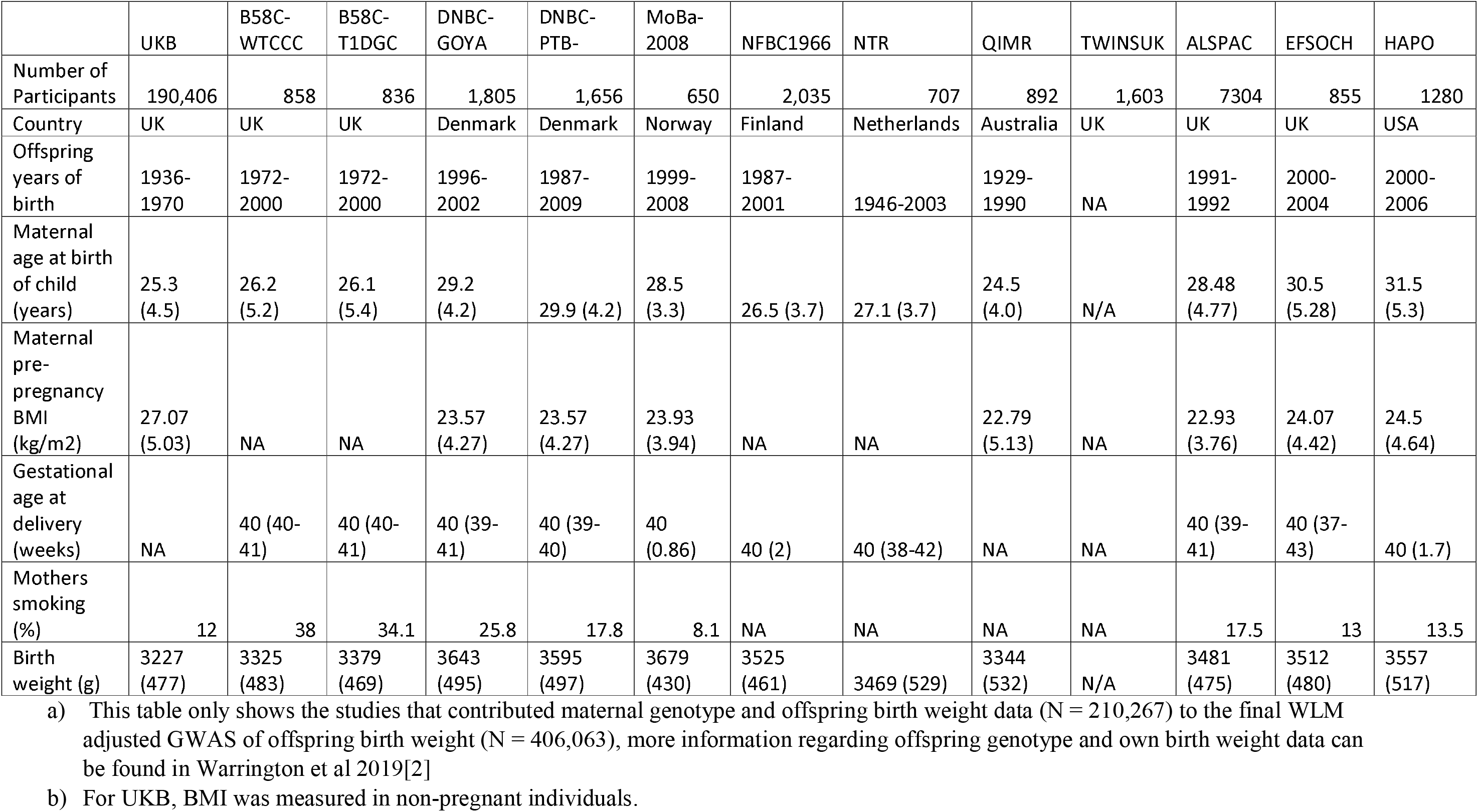
Characteristics of studies contributing maternal genotype and offspring birth weight data to the GWAS of offspring birth weight (data from Warrington et al 2019[2] and Beaumont et al 2018[39])

**Table 2:** Characteristics of the studies used for the secondary analyses.

|  | ALSPAC | BiB | EFSOCH | HAPO 1<sup>a</sup> | HAPO 2<sup>a</sup> |
| --- | --- | --- | --- | --- | --- |
| Number of Participants | 7411 | 3308 | 1022 | 1052 | 815 |
| Country | United Kingdom | United Kingdom | United Kingdom | United States | United States |
| Offspring years of birth | 1991-1993 | 2007-2011 | 2000-2004 | 2001-2006 | 2000-2006 |
| Maternal Age at birth of child (years) | 28.5 (4.8) | 27.1 (6) | 30.4 (5.3) | 32.1 (5.1) | 29.9 (5.4) |
| Maternal pre-pregnancy BMI (kg/m<sup>2</sup>) | 22.9 (3.8) | 26.6 (5.9) | 24 (4.4) | 24.2 (4.6) | 24.6 (5.3) |
| Gestational age at delivery (weeks) | 39.6 (1.7) | 39.7 (1.8) | 39.9 (1.5) | 40 (1.2) | 40 (1.2) |
| Offspring sex (% male) | 49.8 | 51.6 | 51.6 | 47.9 | 50.9 |
| Mothers smoking (%) | 17.2 | 33.1 | 13.3 | 12.9 | 15.1 |
| Birth Weight (g) | 3495 (470.6) | 3438.9 (481.8) | 3513.2 (475.5) | 3542.5 (509.1) | 3539.5 (431.1) |
| Birth Length (cm) | 50.9 (2.2) | NA | 50.3 (2.1) | 50.5 (2.2) | 51.8 (2.5) |
| Birth Ponderal Index (kg/m<sup>3</sup>) | 26.4 (2.7) | NA | 27.7 (2.6) | 27.4 (3.3) | 25.4 (3.3) |
| Birth Head Circumference (cm) | 35 (1.4) | 34.7 (1.4) | 35.2 (1.3) | 34.9 (1.6) | 34.9 (1.4) |
| Birth Triceps Skinfolds (mm) | NA | 5.2 (1.1) | 4.9 (1.1) | 4.1 (0.8) | 4.1 (0.9) |
| Birth Subscapular Skinfolds (mm) | NA | 4.9 (1.1) | 4.9 (1.2) | 4.6 (1) | 4.3 (1) |
| Sum of Birth Skinfolds (mm) | NA | 10.1 (2.1) | 9.7 (2.1) | 13.1 (2.5) | 12.3 (2.4) |
| Cord-blood C-Peptide (µg/mL)<sup>b</sup> | NA | NA | NA | 0.9 (0.7-1.3) | 0.9 (0.7-1.3) |
| Cord-blood insulin (pmol/mL)<sup>b</sup> | NA | 24.3 (15-41) | 37.6 (26-60) | NA | NA |
| Cord-blood leptin (ng/mL)<sup>b</sup> | NA | 7.3 (4-13.1) | NA | NA | NA |
| Cord-blood adiponectin (µg/ml)<sup>b</sup> | NA | 33.3 (26.3-42.7) | NA | NA | NA |
| Fasting Glucose (mmol/l) | NA | 4.4 (0.42) | 4.35 (0.38) | 4.58 (0.37) | 4.51 (0.34) |
| 2 hour postload glucose (mmol/l) | NA | 5.43 (1.3) | NA | 6.02 (1.2) | 6.06 (1.19) |

a) For HAPO 1, genetic data was stored and analysed at the University Feinberg School of Medicine, Chicago. For HAPO 2, genetic data was stored and analysed at the University of Exeter. These were non-overlapping samples of European mothers and babies.
b) For the cord-blood outcomes, because they have a non-standard distribution, the median and interquartile range are displayed.

### Primary Study: Mendelian Randomisation to test the effect of maternal metabolically favourable adiposity vs general adiposity on offspring birth weight

#### Exposures: sample 1

Metabolically favourable adiposity SNPs were identified from the most recent GWAS (N=442,278)[9]. The GWAS of metabolically favourable adiposity uses a composite phenotype characterized by increased body fat percentage and a metabolic profile related to a lower risk of type 2 diabetes, hypertension and heart disease (see **<u>supplementary methods</u>** and reference[9] for more details); 14 SNPs associated with higher body fat percentage and a “favourable” metabolic profile were identified at *p* < 5 x 10^-8^ and replicated[9]. To facilitate the interpretation of our results we weighted these SNPs by the effect estimates of their association with body fat percentage using the latest GWAS of body fat percentage[11].

We used 76 BMI SNPs as instruments for general adiposity from the most recent GWAS of BMI that did not include the UKB sample GWAS (N=322,154), with their weights being extracted from the same GWAS[12]. We did not use the more recent GWAS that did include UKB[13] as that would result in an overlap between sample 1 (genetic instruments-BMI) and sample 2 (genetic instruments-birth weight), which can result in over fitting of the data, and bias towards confounded associations, if there is weak instrument bias[14]

Further details of the metabolically favourable adiposity and BMI GWAS are provided in **<u>sTable 1</u>**, and the characteristics of the SNPs used in our MR analyses are provided in **<u>sTable 2</u>**.

#### Outcome: sample 2

For our second sample we used the latest GWAS of offspring birth weight from the Early Growth Genetics (EGG) consortium. There was a total of 406,063 participants who contributed to the weighted linear model analyses (WLM, see below), of which 101,541 were UK Biobank (UKB) participants who reported their own birth weight and birth weight of their first child, 195,815 UKB and EGG participants with own birth weight data, and 108,707 UKB and EGG participants with offspring birth weight data (see **<u>supplementary methods</u>**)[2].

Birth weight was standardized within UKB and each of the EGG cohorts so that our measurements are in SD units (1 SD of birthweight ≈ 484g, the average SD for birth weight in 18 studies in an early birth weight GWAS[15]). To ensure our analyses considered only the effect of the maternal genotype, and not the correlated fetal genotype, we used the maternal genetic effect on offspring birth weight that had been estimated using a WLM[2]. WLM is a linear approximation of a structural equation model (SEM) that was developed to combine data from disparate study designs to estimate independent maternal and fetal genetic effects, equivalent to conditional analysis in mother-child pairs. The WLM/SEM method combines studies with own genotype data available in addition to own birth weight and offspring birth weight data, with data from studies with only their own birth weight or offspring birth weight (see **<u>supplementary methods</u>** and references[2, 16] for more details). To confirm we obtained similar causal effect estimates with both the WLM and SEM adjusted summary statistics for birth weight, we applied the SEM method to obtain the maternal specific genetic effect on offspring birth weight at each of the SNPs, adjusted for the fetal genotype, using UKB participants only (own birth weight N = 211,815; offspring birth weight N = 187,120) and repeated the main MR analysis.

#### Main Mendelian Randomisation analysis and sensitivity analyses

We performed two-sample MR using Wald ratios[17], which were calculated by dividing each SNP’s effect on offspring birth weight (maternal genetic effect estimated using WLM) by the same SNP’s effect on the exposure (maternal metabolically favourable adiposity or BMI). Standard errors were calculated by dividing the standard error of the SNP’s effect on offspring birth weight by each SNP’s effect on the exposure. Standard MR methods, such as fixed effect pooled Wald ratios, assumes that genetic influences i) robustly (replicated) and statistically strongly related to the exposure (i.e. metabolically favourable adiposity and BMI), ii) are not related to confounders of exposure on the outcome and iii) only influence the outcome (i.e. birth weight) through the exposure. The third assumption can be violated by horizontal pleiotropy and between SNP Wald ratio heterogeneity is indicative of that. Therefore we used Cochran’s Q test, I^2^ and leave-one-out analysis to explore between SNP heterogeneity[18]. We also used MR-Egger[19], weighted-median estimator[20] and Radial MR[21] as sensitivity analyses to explore the extent horizontal pleiotropy may have biased the results (see **<u>supplementary methods</u>** for further details of these methods and their assumptions).

The resulting effect estimates from our MR analyses are reported as the mean difference in offspring birth weight per 1 SD higher maternal body fat percentage (1 SD of body fat percentage = 6.5%; see **<u>supplementary methods</u>**) for metabolically favourable adiposity, and the mean difference in offspring birth weight per 1 standard deviation increase in maternal BMI (1 SD of BMI = 4 kg/m^2^ [3]) for BMI.

### Secondary study: exploring the potential effects of maternal metabolically favourable adiposity vs. general adiposity (BMI) on maternal glucose, cord insulin and neonatal anthropometric traits

We undertook exploratory studies of the effects of metabolically favourable adiposity and BMI on fasting glucose and postload glucose in the general population and in pregnancy, the fetal insulin response to changes in circulating glucose levels (assessed using cord-blood insulin, c-peptide or cord-blood adiponectin) and additional (to birth weight) infant measurements of anthropometry, including head circumference, body length, ponderal index, and triceps and subscapular skinfolds, as well as cord-blood leptin as a marker of total fat mass. Individual participant data from four birth cohorts contributed to one or more of the analyses with these outcomes. Although we made every effort to bring together all relevant studies with maternal and offspring genotype and these outcomes, these analyses are considered exploratory analyses because of the relatively small sample sizes for MR studies that we were able to obtain. As these were exploratory analyses we undertook main analyses using the Wald ratio method (see above) but did not undertake MR sensitivity analyses, such as MR-Egger, to explore horizontal pleiotropy.

#### Contributing cohorts

The four birth cohorts we used to perform these exploratory analyses were the Avon Longitudinal Study of Parents and Children (ALSPAC)[22, 23], Born in Bradford (BiB)[24], the Exeter Family Study of Childhood Health (EFSOCH)[25] and Hyperglycemia and Adverse Pregnancy Outcomes (HAPO)[26] (maximum N = 9,323 mother-child pairs).

Further information on these cohorts and their contribution to the study can be found in **<u>Table 2</u>** and **<u>sFigure 1</u>**. See **<u>supplementary methods</u>** for the cohort descriptions and for additional information on the data extraction, genotyping and phenotype assessment as well as how ethnicity was defined for each study.

#### Mendelian Randomisation analysis and sensitivity analysis

We used fixed effect pooled Wald ratio analysis to determine the effect of maternal metabolically favourable adiposity and BMI on offspring birth weight in each of these studies and compared the pooled result of that with the same result in our main analyses of effects on birth weight. The maternal genotype effects on offspring birth weight were directly adjusted for the fetal genotype, this being done to avoid violating the third assumption of MR analyses (that the instrument only influences the outcome via the exposure).

In the absence of any publically available GWAS of fasting glucose in pregnancy, we used published GWAS summary statistics from the Meta-Analyses of Glucose and Insulin-related traits Consortium (MAGIC) to investigate the effects of the metabolically favourable adiposity and BMI SNPs on fasting glucose. The MAGIC consortium GWAS reported data from 46,186 White-European (non-pregnant and including women and men) adults on fasting glucose from 17 population cohorts and four case control studies in the discovery data set (there were 122,743 adults in the total data set)[27]. We checked the consistency of these associations with those for pregnancy fasting glucose in BiB, EFSOCH and HAPO (no pregnancy fasting glucose data was available for ALSPAC).

### Assessing the plausibility of MR assumptions for primary and secondary studies

Sensitivity analyses for assessing the plausibility of the assumption that the genetic instruments are not related to other factors (i.e. through horizontal pleiotropy) that also influence birth weight are described above for the primary study. Full details of how we explored the validity of the genetic instrumental variables and other potential violations of MR assumptions in both the primary and secondary study are provided in the **<u>supplementary methods</u>**.

## Results

The associations between the SNPs analysed in the primary study and UKB+EGG birth weight are shown in **<u>sTables 3</u>** and the association between the same SNPs and all the outcomes analysed for the secondary study (fasting glucose, pregnancy fasting glucose, pregnancy two hour glucose, birth weight, birth length, ponderal index, head circumference, triceps skinfold thickness, subscapular skinfold thickness, sum of skinfold thickness, cord-insulin, cord-c-peptide, cord-leptin and cord-adiponectin) are shown in **<u>sTables 4, 5 and 6</u>**.

### Maternal metabolically favourable adiposity and maternal general adiposity, indexed by BMI, have opposite effects on offspring birth weight

We found evidence that higher maternal metabolically favourable adiposity causes lower offspring birth weight (**<u>Figure 2</u>**). The main estimate (−94g (95% CI, −150 to −38) of difference in mean birth weight per 1 SD (6.5 %) higher maternal metabolically favourable body fat percentage) was consistent with both the MR-Egger and weighted-median estimates (**<u>Figure 2</u>**).There was evidence of heterogeneity between the Wald ratios across the SNPs (Cochrans Q = 33.46 (d.f. = 13), I^2^ = 61.1 %), yet results were consistent across leave-one-out analysis (**<u>sFigure 2</u>**). Using the SEM to adjust for fetal genotype effects also gave very similar results (**<u>sFigure 3</u>**).

**Figure 2:**
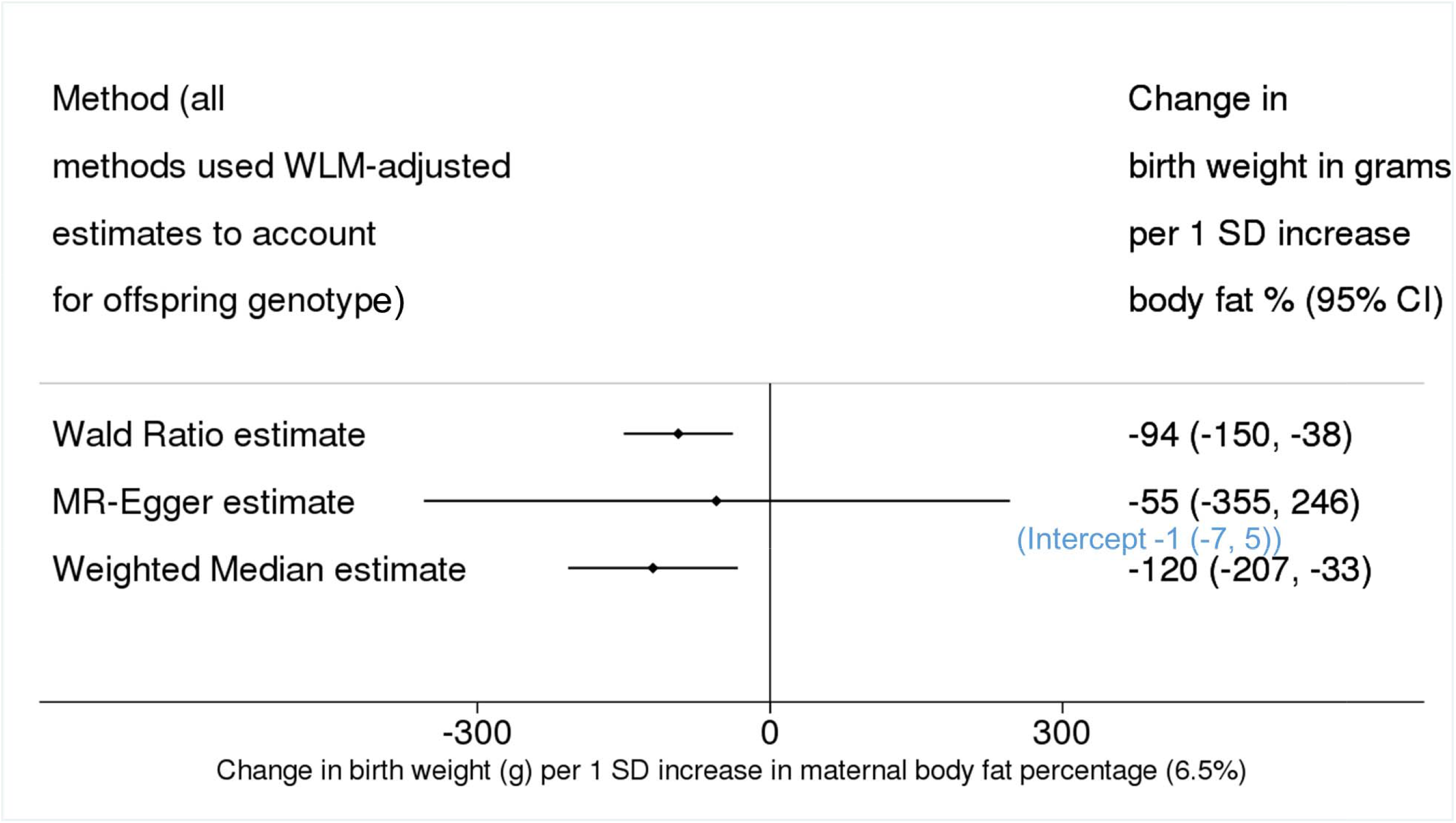
Causative effect estimates for maternal metabolically favourable adiposity on offspring birth weight

The two-sample MR estimates for BMI are consistent with higher maternal general adiposity leading to increased offspring birth weight (**<u>Figure 3</u>**). The main MR estimate was 35g (95% CI, 16 to 53) of difference in mean birth weight per 1 SD (4 kg/m^2^) higher maternal BMI. MR-Egger (20g (95% CI, −51 to 92)) and weighted-median (14g (95% CI, −20 to 48)) estimates were directionally the same, though smaller than the main estimate (**<u>Figure 3</u>**).In this analysis, there was also evidence of heterogeneity between the Wald ratios for each SNP (Cochrans Q = 178.42 (d.f. = 75), I^2^ = 58 %), yet again results were consistent across leave-one-out analysis (**<u>sFigure 4</u>**). Using the SEM to adjust for fetal genotype gave very similar results (**<u>sFigure 5</u>**).

**Figure 3:**
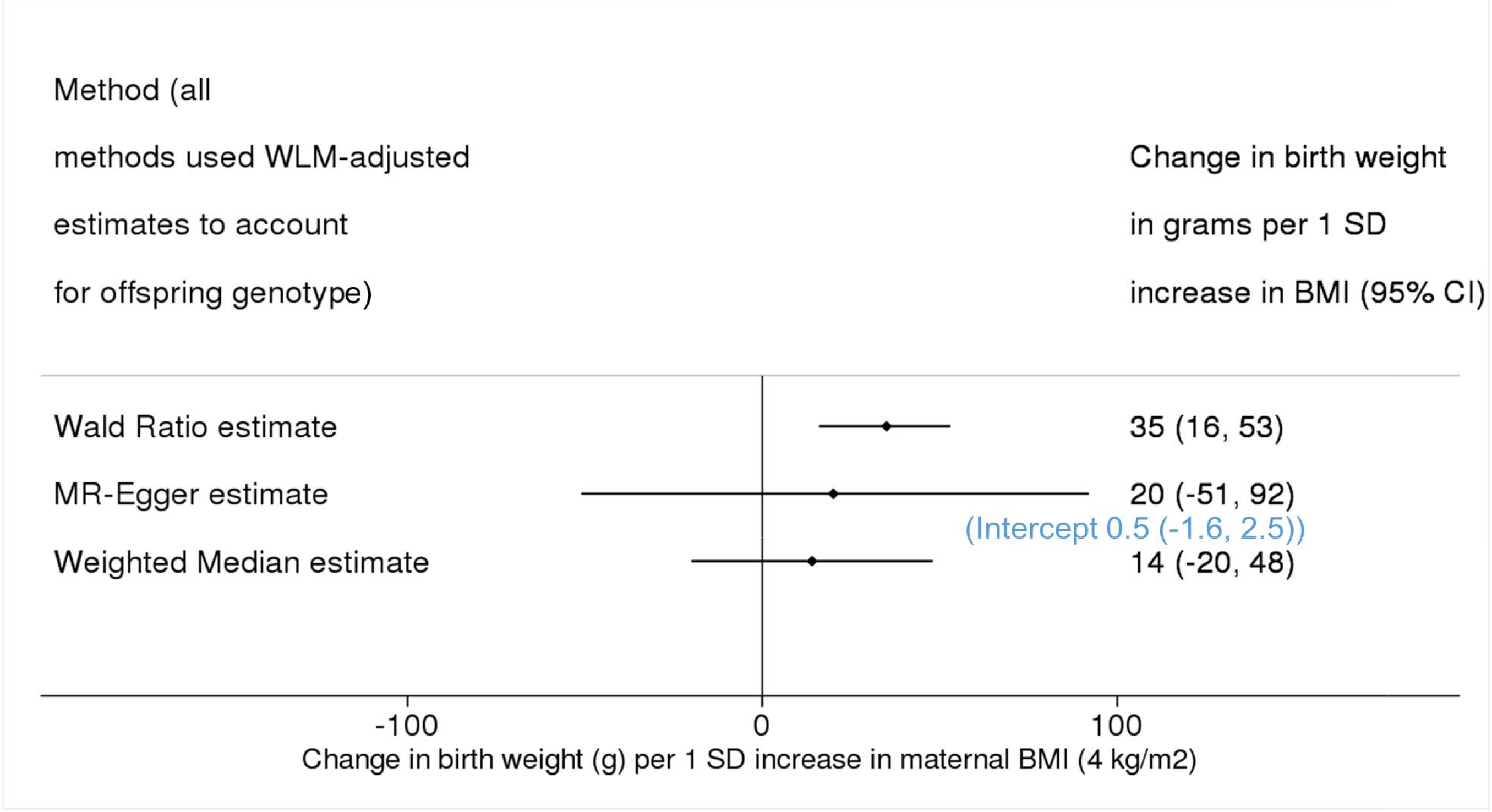
Causative effect estimates for maternal BMI on offspring birth weight

We used the radial MR-Egger method to detect the presence and impact of outlier SNPs. The radial MR-Egger estimate for the effect of maternal metabolically favourable adiposity on offspring birth weight was close to null (−2g (95% CI, −184 to 180)) (**<u>sFigure 6 and sFigure 7</u>**), and four MR-Egger outliers were identified. In analysis with these removed the effect estimate was stronger than in the main analysis (−248g, (95% CI, −490 to −6)) (**<u>sFigure 6</u>**).

The radial MR-Egger estimate of the effect of maternal BMI on offspring birth weight was weaker (13g (95% CI, −37 to 64), see **<u>sFigure 8 and sFigure9</u>**) compared to the main estimate, and 18 outliers were identified. In analysis with these removed the effect estimate became more consistent (22g (95% CI, −32 to 77)) with the main result (**<u>sFigure 8</u>**).

### Secondary study results

Using published data from MAGIC we showed that higher metabolically favourable adiposity resulted in lower fasting glucose in men and non-pregnant women, whilst higher maternal BMI resulted in higher fasting glucose (**<u>Figure 4</u>**). The relationship between maternal metabolically favourable adiposity, or BMI, and fasting glucose in pregnancy were consistent with this, as was 2 hour post-prandial glucose in pregnancy (**<u>Figure 4</u>**). The point estimate for the effect of metabolically favourable adiposity on 2 hour post prandial glucose was greater than that for fasting glucose, but confidence intervals were wider (**<u>Figure 4</u>**).

**Figure 4:**
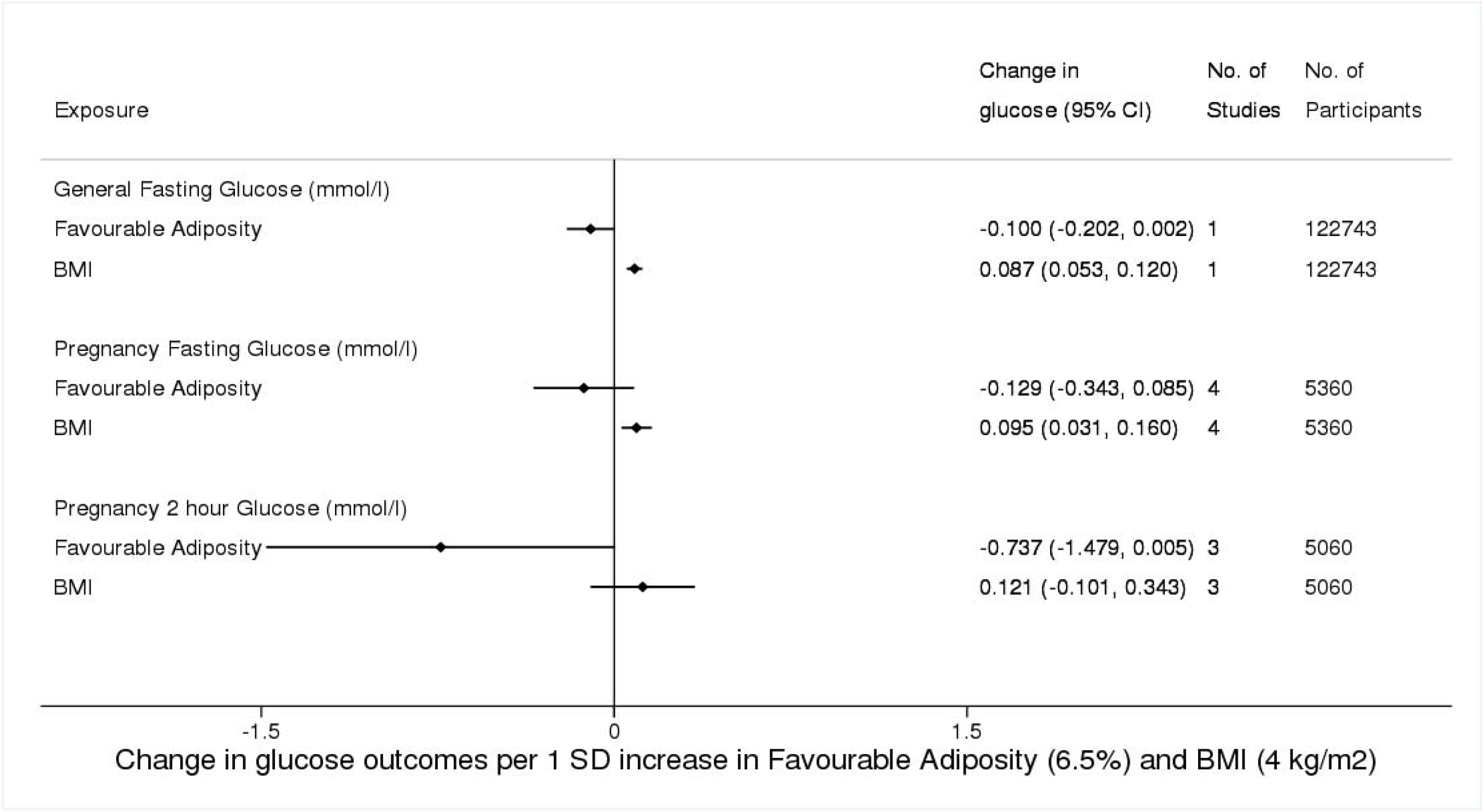
Causative effect estimates for maternal BMI and metabolically favourable adiposity on maternal pregnancy glucose outcomes

Evidence suggested that maternal metabolically favourable adiposity consistently resulted in lower neonatal anthropometric outcomes (**<u>Figure 5</u>**). In contrast, evidence suggested that maternal BMI consistently resulted in higher neonatal anthropometric outcomes (**<u>Figure 5</u>**).

Higher maternal metabolically favourable adiposity showed suggestive evidence of causing lower cord-blood leptin levels, in contrast there was strong evidence of higher maternal BMI resulting in higher cord-blood leptin levels (**<u>Figure 6</u>**). There was no detectable effect of maternal metabolically favourable adiposity or BMI on cord-blood insulin, c-peptide, or adiponectin levels, though the small sample sizes made these results imprecise (**<u>Figure 6</u>**).

**Figure 5:**
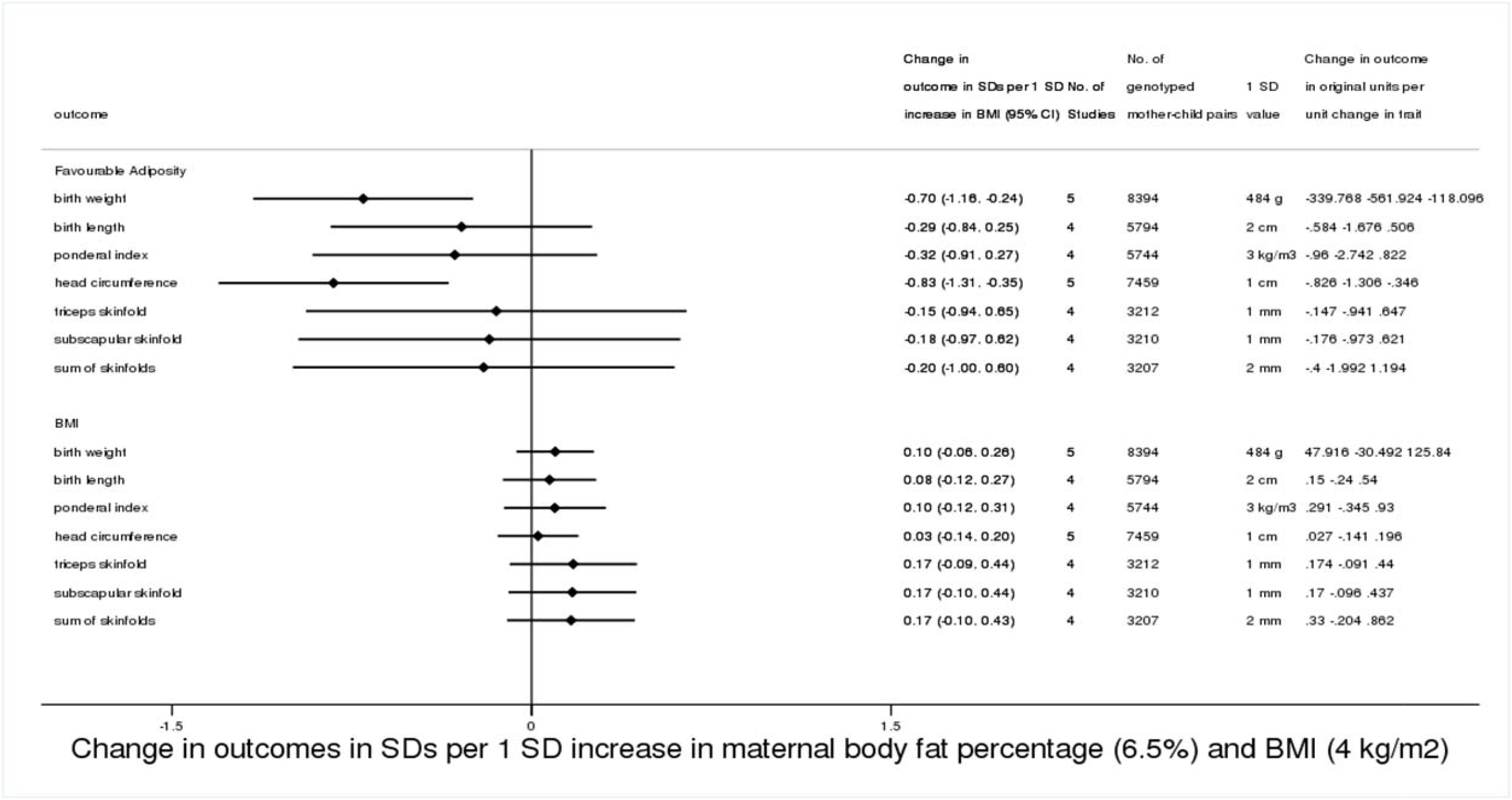
Causative effect estimates for maternal BMI and metabolically favourable adiposity on other birth anthropometric outcomes, adjusted for offspring genotype

**Figure 6:**
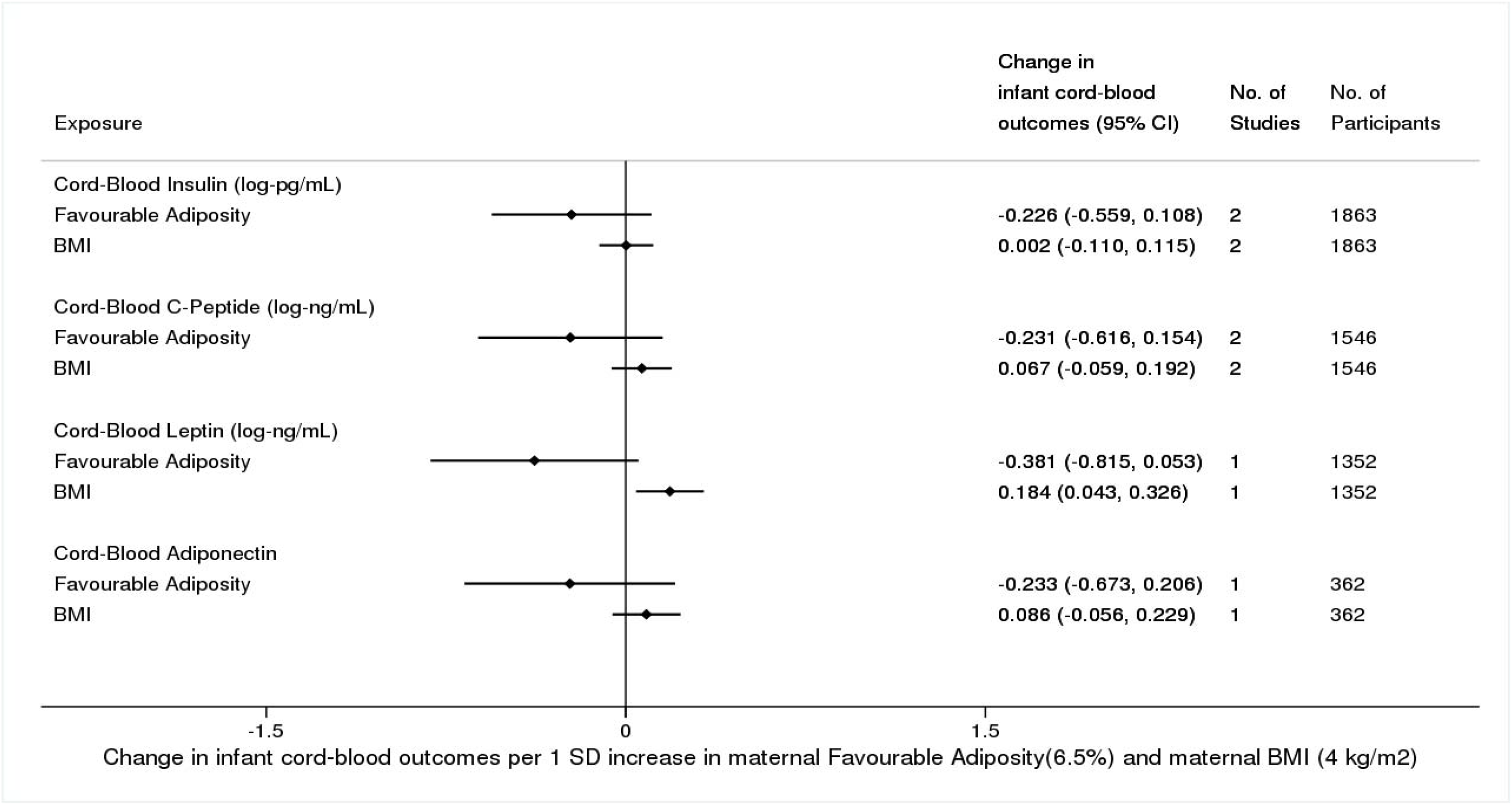
Causative effect estimates for maternal BMI and metabolically favourable adiposity on infant cord-blood outcomes, adjusted for offspring genotype

### Validity of the genetic instrumental variables

The BMI SNPs were positively and consistently associated with pregnancy BMI across all of the cohorts used for our secondary analyses. There was no evidence that collider bias influenced the results of maternal metabolically favourable adiposity on outcomes (see **<u>supplementary methods</u>**).

The metabolically favourable adiposity genetic score was not associated with maternal smoking or maternal education level in UKB, ALSPAC, EFSOCH or BiB, but was associated with lower female educational level in HAPO (**<u>sTable 8</u>**). The BMI genetic score was associated with an increased prevalence of current smoking status in women in UKB and pregnancy smoking status in ALSPAC and with lower female educational level in UKB (**<u>sTable 8</u>**). Given these findings, we undertook further analyses using multivariable MR[28] to see whether maternal education and smoking status confounded the association (for more details see **<u>supplementary methods</u>**). Results from the multivariable adjusted MR analyses were consistent with those from the unadjusted MR analyses, for both smoking and years in education, adjusted for individually and when both were included in the multivariable model (**<u>sTable 9</u>**).

## Discussion

This study applied Mendelian Randomization to compare the effects of maternal favourable adiposity with general adiposity (BMI) on offspring birth weight. We found evidence of contrasting effects; while higher maternal favourable adiposity was related to lower offspring birth weight, the opposite was the case for maternal general adiposity. Our secondary analyses provided some evidence that higher maternal favourable adiposity causes lower fasting glucose both inside and outside of pregnancy, i.e. has the opposite effect to that of higher BMI. Additional analyses of effects of maternal favourable vs. general adiposity on different neonatal anthropometric outcomes were consistent in direction with those of birth weight, but there was insufficient power to determine whether effect sizes varied between outcomes that capture fat mass (e.g. skinfolds) vs. lean mass (e.g. length). There was also evidence to suggest that higher maternal favourable adiposity causes lower cord-blood leptin levels whilst higher maternal BMI causes higher cord-blood leptin levels. As cord-blood leptin is a biomarker for fetal fat-mass this provides some support that higher maternal metabolically favourable adiposity might result in lower fetal fat accumulation while higher maternal BMI results in greater fetal fat accumulation. There was insufficient power to detect precise effects on cord-blood markers of fetal insulin response as studies used different measures of this (insulin, c-peptide or adiponectin).

The causal effects of metabolically favourable adiposity have previously been investigated for other health outcomes, such as type II diabetes, hypertension, heart disease, fatty liver and depression in adults[9, 29, 30]. With the exception of depression (where both higher BMI and metabolically favourable adiposity were associated with higher risk of depression[30]), metabolically favourable adiposity has been found to have the opposite causal effect to BMI[9, 29]. Thus our novel finding of higher maternal metabolically favourable adiposity being associated with lower offspring birth weight has consistency with this emerging literature on other adult outcomes.

Metabolically favourable adiposity is a composite of individual traits, many of which have been found to be causally associated with birth weight. In particular, higher maternal fasting glucose (often resulting from insulin resistance, a component of the metabolically favourable adiposity trait) has consistently been found to be causally associated with higher offspring birth weight in MR studies[2, 3]. We have shown higher metabolically favourable adiposity to be tentatively associated with lower fasting glucose, both outside and during pregnancy, suggesting that the effect of higher maternal metabolically favourable adiposity on lower offspring birth weight may be mediated by its effect on fasting glucose levels. Though we have focussed on fasting glucose, metabolically favourable adiposity is also associated with other exposures, in particular lower triglyceride levels[9]. However, MR studies have failed to find any evidence of an association between maternal circulating triglyceride levels and offspring birth weight[3, 31], furthermore glucose has been shown to cross the placenta[32] (thus be able to directly influence the fetus), for that reason our focus in this study was on maternal fasting glucose.

### Study Strengths and Limitations

To the best of our knowledge, this is the first study to use MR to investigate the effect of maternal metabolically favourable adiposity on offspring birth weight. We used data from a very large GWAS of birth weight and also for the first time examined potential effects on maternal glucose traits, additional (to birth weight) infant anthropometric measurements and cord-blood measurements in exploratory analyses. We have explored the validity of our genetic instrumental variables using multiple sensitivity analyses, including the recently developed Radial MR method[21], and found that overall results from these sensitivity analyses were consistent with our main findings. Our BMI genetic variants explained 2.7% of the variance in BMI[12]. As metabolically favourable adiposity is not a directly measured single trait, it is not possible to measure how much “variance” is explained by the genetic variants. However for the primary study with birth weight the MR estimates were precise.

A potential limiting factor may be the low response rate for UKB and the maternal report of offspring birth weight years after the birth in that study. Research suggests that a highly select cohort (as in the case of UKB with a 5% response[33]) can result in selection bias in genetic or MR analyses[34, 35]. The fact that the birth weight results from the secondary study, using four birth cohorts with response rates of at least 70%, were consistent in magnitude and direction with the primary study suggests this is unlikely to be a major source of bias. However, three of the four cohorts used (ALSPAC, EFSOCH and HAPO) were also used in the EGG consortium GWAS of birth weight and thus are not independent of the main result. Self-report of own and first child’s birth weight and the rounding to the nearest 1 pound (~0.454kg, first child’s birth weight only) may have introduced error in the birth weight measure in UKB, but this would be random with respect to genotype and expected to bias results towards the null. In UKB there is a relatively lower reported birth weight than in most of the other cohorts used in this study, which is likely to reflect secular trends of increasing birth size over time, given that the women in UKB were born between the early 1940s and early 1960s, whereas the majority of the participants in the other EGG cohorts were born after the mid-1980s.

In UKB, genetically instrumented BMI was found to be associated with both educational attainment and with smoking status. Smoking has been shown to correlate with lower birth weight, and educational attainment is a marker of socioeconomic position, hence both these traits could result in violation of the assumption that the genetic instruments do not relate to other (than the exposure of interest) risk factors for the outcome[36]. However, results of the multivariable IVW analyses of maternal BMI on offspring birth weight adjusted for smoking and years in education were not substantially different from the main result, suggesting the association was not heavily confounded.

Whilst there was evidence of between SNP heterogeneity of both metabolically favourable adiposity and BMI, results were directionally consistent across the main estimate, leave-one-out analyses, MR-Egger and weighted-median analyses. The Radial MR analyses for both maternal metabolically favourable adiposity and maternal BMI found evidence of SNPs with outlier effects (four for maternal metabolically favourable adiposity, 18 for maternal BMI), suggesting that pleiotropic effects may bias the main estimates. However, in both cases, removal of the outlier SNPs from the analyses resulted estimates consistent with the main estimates, suggesting horizontal pleiotropy is unlikely to be a major source of bias for our analyses. In our secondary studies we had insufficient power to undertake these sensitivity analyses.

Our analyses hinged on using various methods (including WLM, SEM and mother-child pair analyses) to adjust for fetal effects on offspring birth weight. Adjustment for fetal genotype introduces a spurious association between maternal and paternal genotype due to collider bias effects. Nonetheless, it seems mechanistically unlikely that adiposity of the father could directly (i.e. not via offspring genetic inheritance of paternal genetic variants) influence offspring birth weight and in previous sensitivity analyses it has been shown that any bias from not being able to adjust for the paternal genotype is small in comparison to the bias that would be introduced if the fetal genotype is not controlled for[37, 38].

In conclusion, our results suggest that maternal metabolically favourable adiposity has the opposite effect on offspring birth weight to that of maternal BMI. This means that higher adiposity in mothers does not necessarily lead to higher offspring birth weight, and may result in lower offspring birth weight if accompanied by a favourable metabolic profile. In the future, methods to stratify overweight and obese pregnant women by their metabolically favourable adiposity status could allow for targeted interventions to achieve healthy birth weight.

## Data Availability

Our study uses two-sample Mendelian randomization (MR). We used both published summary results (i.e. taking results from published research papers and websites) and individual participant cohort data as follows:
Journal published and website summary data were used for sample one of the two sample Mendelian randomization (published GWAS of BMI and body fat percentage). The references to those published data sources are provided in the main paper. The data for the GWAS of BMI is available here.
https://portals.broadinstitute.org/collaboration/giant/index.php/GIANT_consortium_data_files
The data for the GWAS of body fat percentage is available here.
https://walker05.u.hpc.mssm.edu
We used individual participant data for the second MR sample and for undertaking sensitivity analyse from the UKB, ALSPAC, BiB, EFSOCH and HAPO cohorts.
The data in UKB, ALSPAC and BiB are fully available, via managed systems, to any researchers. The managed system for both studies is a requirement of the study funders but access is not restricted on the basis of overlap with other applications to use the data or on the basis of peer review of the proposed science. Researchers have to pay for a dataset to be prepared for them.
UKB. Full information on how to access these data can be found here - https://www.ukbiobank.ac.uk/using-the-resource/
ALSPAC. The ALSPAC data management plan (http://www.bristol.ac.uk/alspac/researchers/data-access/documents/alspac-data-management-plan.pdf) describes in detail the policy regarding data sharing, which is through a system of managed open access. The steps below highlight how to apply for access to the data included in this paper and all other ALSPAC data.
1. Please read the ALSPAC access policy (PDF, 627kB) which describes the process of accessing the data and samples in detail, and outlines the costs associated with doing so.
2. You may also find it useful to browse the fully searchable ALSPAC research proposals database, which lists all research projects that have been approved since April 2011.
3. Please submit your research proposal for consideration by the ALSPAC Executive Committee. You will receive a response within 10 working days to advise you whether your proposal has been approved.
If you have any questions about accessing data, please.
BiB. Full information on how to access these data can be found here - https://borninbradford.nhs.uk/research/how-to-access-data/
EFSOCH. Requests for access to the original EFSOCH dataset should be made in writing in the first instance to the EFSOCH data team via the Exeter Clinical Research Facility.

https://portals.broadinstitute.org/collaboration/giant/index.php/GIANT_consortium_data_files

https://walker05.u.hpc.mssm.edu

https://www.ukbiobank.ac.uk/using-the-resource/

http://www.bristol.ac.uk/alspac/researchers/data-access/documents/alspac-data-management-plan.pdf

https://borninbradford.nhs.uk/research/how-to-access-data/

## Acknowledgements

This research has been conducted using the UKB Resource under application number 7036. We would like to thank the participants and researchers from the UKB who contributed or collected data. We are extremely grateful to all of the families who took part in ALSPAC, the midwives for their help in recruiting them, and the whole ALSPAC team, which includes interviewers, computer and laboratory technicians, clerical workers, research scientists, volunteers, managers, receptionists and nurses. BiB is only possible because of the enthusiasm and commitment of the children and parents. We are grateful to all the participants, practitioners, and researchers who have made BiB happen. We are also grateful for the families that took part in EFSOCH and the researchers that collected data. We are grateful to the Genetics of Complex traits team at the University of Exeter, for their assistance in learning the methods and navigating the study data; in particular we are grateful to Francesco Casanova who helped with the data extraction for BiB. The authors would like to acknowledge the use of the University of Exeter High-Performance Computing (HPC) facility in carrying out this work.

## Data Availability

Our study uses two-sample Mendelian randomization (MR). We used both published summary results (i.e. taking results from published research papers and websites) and individual participant cohort data as follows:

Journal published and website summary data were used for sample one of the two sample Mendelian randomization (published GWAS of BMI and body fat percentage). The references to those published data sources are provided in the main paper. The data for the GWAS of BMI is available here.

https://portals.broadinstitute.org/collaboration/giant/index.php/GIANT_consortium_data_files

The data for the GWAS of body fat percentage is available here.

https://walker05.u.hpc.mssm.edu

We used individual participant data for the second MR sample and for undertaking sensitivity analyse from the UKB, ALSPAC, BiB, EFSOCH and HAPO cohorts.

The data in UKB, ALSPAC and BiB are fully available, via managed systems, to any researchers. The managed system for both studies is a requirement of the study funders but access is not restricted on the basis of overlap with other applications to use the data or on the basis of peer review of the proposed science. Researchers have to pay for a dataset to be prepared for them.

UKB. Full information on how to access these data can be found here - https://www.ukbiobank.ac.uk/using-the-resource/

ALSPAC. The ALSPAC data management plan

(http://www.bristol.ac.uk/alspac/researchers/data-access/documents/alspac-data-management-plan.pdf) describes in detail the policy regarding data sharing, which is through a system of managed open access. The steps below highlight how to apply for access to the data included in this paper and all other ALSPAC data.

1. Please read the ALSPAC access policy (PDF, 627kB) which describes the process of accessing the data and samples in detail, and outlines the costs associated with doing so.
2. You may also find it useful to browse the fully searchable ALSPAC research proposals database, which lists all research projects that have been approved since April 2011.
3. Please submit your research proposal for consideration by the ALSPAC Executive Committee. You will receive a response within 10 working days to advise you whether your proposal has been approved.

If you have any questions about accessing data, please.

BiB. Full information on how to access these data can be found here - https://borninbradford.nhs.uk/research/how-to-access-data/

EFSOCH. Requests for access to the original EFSOCH dataset should be made in writing in the first instance to the EFSOCH data team via the Exeter Clinical Research Facility.

## Author Contributions

DAL and RMF designed this study, with WDT further developing the design.

DAL (ALSPAC), ATH and BAK (EFSOCH), DS and WLL (HAPO) and GS, RA, DM and DAL (BiB) contributed to the collection of and management of cohort data.

ARW, RNB and JT prepared the imputed genotype and phenotype data for the UKB sample of European ancestry participants.

TMF, HY and YJ performed a multivariable GWAS to discover favourable adiposity genetic variants.

NMW and DME performed a Weighted Linear Model GWAS using the UKB + EGG genetic and birth weight data.

WDT, RMF, DAL and MC-B wrote the analysis plan, and WDT undertook most of the analyses with support from JT, M-CB, RB, RMF and DAL.

AK carried out data analyses using the HAPO dataset in Chicago, and in this task he was supervised by DS.

WDT wrote the first draft of the paper with support from M-CB, RMF and DAL; all other authors read and made critical revisions to the paper.

WDT, RMF and DAL act as guarantors for the paper’s integrity

**Abbreviations**
ALSPAC: Avon Longitudinal Study of Parents and Children),
BiB: Born in Bradford),
EFSOCH: Exeter Family Study of Childhood Health),
EGG: Early Growth Genetics consortium),
HAPO: Hyperglycaemia and Adverse Pregnancy Outcomes),
MAGIC: Meta-Analysis of Glucose and Insulin related traits Consortium),
MR: Mendelian Randomisation),
UKB: United Kingdom Biobank),
SEM: Structural Equation Modelling),
WLM: (Weighted Linear Modelling)

